# Diagnosing SARS-CoV-2 infection: the danger of over-reliance on positive test results

**DOI:** 10.1101/2020.04.26.20080911

**Authors:** Andrew N. Cohen, Bruce Kessel, Michael G. Milgroom

**Affiliations:** Center for Research on Aquatic Bioinvasions, Richmond CA, USA; John A. Burns School of Medicine, University of Hawai’i, Honolulu HI, USA.; School of Integrative Plant Science, Plant Pathology and Plant-Microbe Biology Section, Cornell University, Ithaca NY, USA.

## Abstract

Contrary to the practice during previous epidemics, with COVID-19 health authorities have treated a single positive result from a PCR-based test as confirmation of infection, irrespective of signs, symptoms and exposure. This is based on a widespread belief that positive results in these tests are highly reliable. However, evidence from external quality assessments and real-world data indicate enough a high enough false positive rate to make positive results highly unreliable over a broad range of scenarios. This has clinical and case management implications, and affects an array of epidemiological statistics, including the asymptomatic ratio, prevalence, and hospitalization and death rates, as well as epidemiologic models. Steps should be taken to raise awareness of false positives and reduce their frequency. The most important immediate action is to check positive results with additional tests, at least when prevalence is low.

**Key messages:** The high specificities (usually 100%) reported in PCR-based tests for SARS-CoV-2 infection do not represent the real-world use of these tests, where contamination and human error produce significant rates of false positives.

Widespread lack of awareness of the real-world false positive rates affects an array of clinical, case management and health policy decisions. Similarly, health authorities’ guidance on interpreting test results is often wrong.

Steps should be taken immediately to reduce the frequency and impacts of false positive results, including checking positive results with additional tests at least when prevalence is low.

---

Most tests for active SARS-CoV-2 infection use the polymerase chain reaction (PCR) to amplify and detect diagnostic sequences within the virus’ RNA. According to leading health authorities, while negative results from these tests are frequently wrong, positive results are highly reliable.^1-4^ Accordingly, the World Health Organization (WHO) and most government health ministries diagnose SARS-CoV-2 infection on the basis of a single positive PCR result, even in asymptomatic persons without any history of exposure.^5-11^ For example, WHO defines a confirmed case as a person with a positive test result, “irrespective of clinical signs and symptoms.”^5^

This is a departure from historical practice. In previous epidemics case definitions required individuals to be symptomatic, and health authorities voiced concerns that false positive results from PCR-based tests could harm both the individuals tested and the ability of agencies to monitor outbreaks. National and international health agencies adopted measures to limit the occurrence of false positives, recommending that PCR-based testing be limited to individuals with a high probability of infection (those with symptoms and/or significant exposure), and often requiring confirmation of positive results by a second, independent test (Box 1). These warnings and requirements are absent from the same agencies’ current guidance on SARS-CoV-2 testing.

## Box 1: Measures minimizing false positive results in PCR-based tests

### Then

#### SARS-CoV-1

**US CDC:** “To decrease the possibility of a false-positive result, testing should be limited to patients with a high index of suspicion for having SARS-CoV disease…In addition, any positive specimen should be retested in a reference laboratory to confirm that the specimen is positive. To be confident that a positive PCR specimen indicates that the patient is infected with SARS-CoV, a second specimen should also be confirmed positive.”^12^

**WHO:** “[Requirements for the laboratory diagnosis of SARS…almost always involves two or more different tests or the same assay on two or more occasions during the course of the illness or from different clinical sites…A single test result is insufficient for the definitive diagnosis of SARS-CoV infection.”^13^

#### H1N1 Influenza Virus

US CDC: Case confirmation requires presentation with an influenza-like illness in addition to a single positive PCR test.^14^

#### MERS-CoV

**US CDC:** Requirements for testing include both specific clinical features and epidemiologic risk,^15^ and positive results must be confirmed by the CDC.^16^

**WHO:** Testing should be limited to persons with specified symptoms and, in most cases, elevated risk of exposure.^17^

#### Ebola Virus

**US CDC:** “CDC recommends that Ebola testing be conducted only for persons who…[have] both consistent signs or symptoms and risk factors…Any presumptive positive Ebola test result must be confirmed at the CDC…CDC considers a single diagnostic test…insufficient for public health decision-making.”^18^

**WHO:** Case confirmation requires specific clinical signs in addition to a single positive PCR test.^19^

#### Zika Virus

**US CDC:** Testing is recommended only for pregnant women with symptoms and recent exposure, or asymptomatic pregnant women with ongoing exposure. “[B]ecause of the potential for false-positive…results, updated recommendations include [PCR] testing of both serum and urine and concurrent Zika virus IgM antibody testing to confirm the diagnosis…with more than one test.”^20^

**WHO:** Testing is recommended only for symptomatic patients.^21^

### Now

#### SARS-CoV-2

Except for validation of a laboratory’s first few results, we found no requirement or recommendation for a second confirmatory test in guidance documents from the World Health Organization, the US Centers for Disease Control and Prevention, the European Centre for Disease Prevention and Control, Public Health England, the Public Health Agency of Canada, the Pan American Health Organization, or South Korea’s Centers for Disease Control and Prevention; instead these entities require only a single positive PCR result to confirm infection in symptomatic or asymptomatic persons.^5-1^ The Chinese Centers for Disease Control and Prevention requires clinical manifestations and usually exposure history in addition to a positive PCR result to confirm a case.^22^ On May 27 the Norwegian Institute of Public Health amended its guidance to recommend confirmatory tests of positive results in persons who are both asymptomatic and without exposure history.^23^

In most regions testing was initially restricted to persons with specified clinical signs and symptoms and exposure history, but as more tests became available many authorities allowed broader use of PCR-based tests, including testing of individuals with no symptoms or known exposure risk.

In this Analysis we argue that basing diagnoses on unrestricted PCR-based testing freed from clinical context has created serious problems. PCR-based tests produce a significant number of false positive results, making positive results unreliable over a broad range of real-world scenarios. Consequently, the frequent assertion that positive test results for SARS-CoV-2 are more reliable than negative results^4^ is wrong most of the time, and the widespread reliance on a single positive PCR result as a sufficient basis for diagnosis has been a mistake. The general misunderstanding of the rate of false positives in SARS-CoV-2 testing affects clinical and case management decisions, and through flawed interpretations of test statistics, has affected policy decisions. As an immediate, minimum step we recommend checking positive PCR results for asymptomatic individuals with a second independent test; over the longer term, we should work on eliminating the underlying causes of false positives.

## False positives

The accuracy of a diagnostic test is measured by sensitivity, which is the proportion of infected individuals that test positive, and specificity, the proportion of uninfected individuals that test negative. Although SARS-CoV-2 PCR assays are widely reported to have 100% specificity^4^— that is, a false positive rate of 0%—this refers only to the tests’ lack of reaction with substances other than SARS-CoV-2 RNA (analytical specificity), and not to the potential for incorrect results in real-world testing (clinical specificity) where contamination and human error can generate false positives during sample collection, transport and analysis.^4^

The only published data on the full false positive rate of SARS-CoV-2 tests in real-world settings appear to be from two studies that found rates of 0.3% and 3% in presurgical patients.^24 25^ Rates within laboratories can be assessed by challenging participating laboratories with prepared samples that either contain or are free of the virus’ RNA. We are aware of seven such assessments, known as external quality assessments or proficiency tests, for SARS-CoV-2.

Four studies tested a total of 119 South Korean laboratories, and reported no positive results for 47, 33, 16 and 236 negative samples.^26 27^ Another study assessed 52 Austrian laboratories, and reported no positive results for 67 negative samples.^28^ The absolute lower detection limit for false positive rates in these studies ranged from 0.4% to 6.3%. A German study of 463 laboratories found an overall false positive rate of 1.9% by gene target, but did not report results for samples.^29^ A study of 365 laboratories in 36 countries reported 11 positive results for 1,529 negative samples, yielding a false positive rate of 0.7%.^30^ These results are generally consistent with data from 43 external quality assessments of similar PCR assays of other RNA viruses conducted in 2004-2019. Out of 10,538 negative samples, 336 (3.2%) were reported as positive. The median false positive rate was 2.3%, and the interquartile range was 0.8-4.0% (Table 1).

**Table 1.**
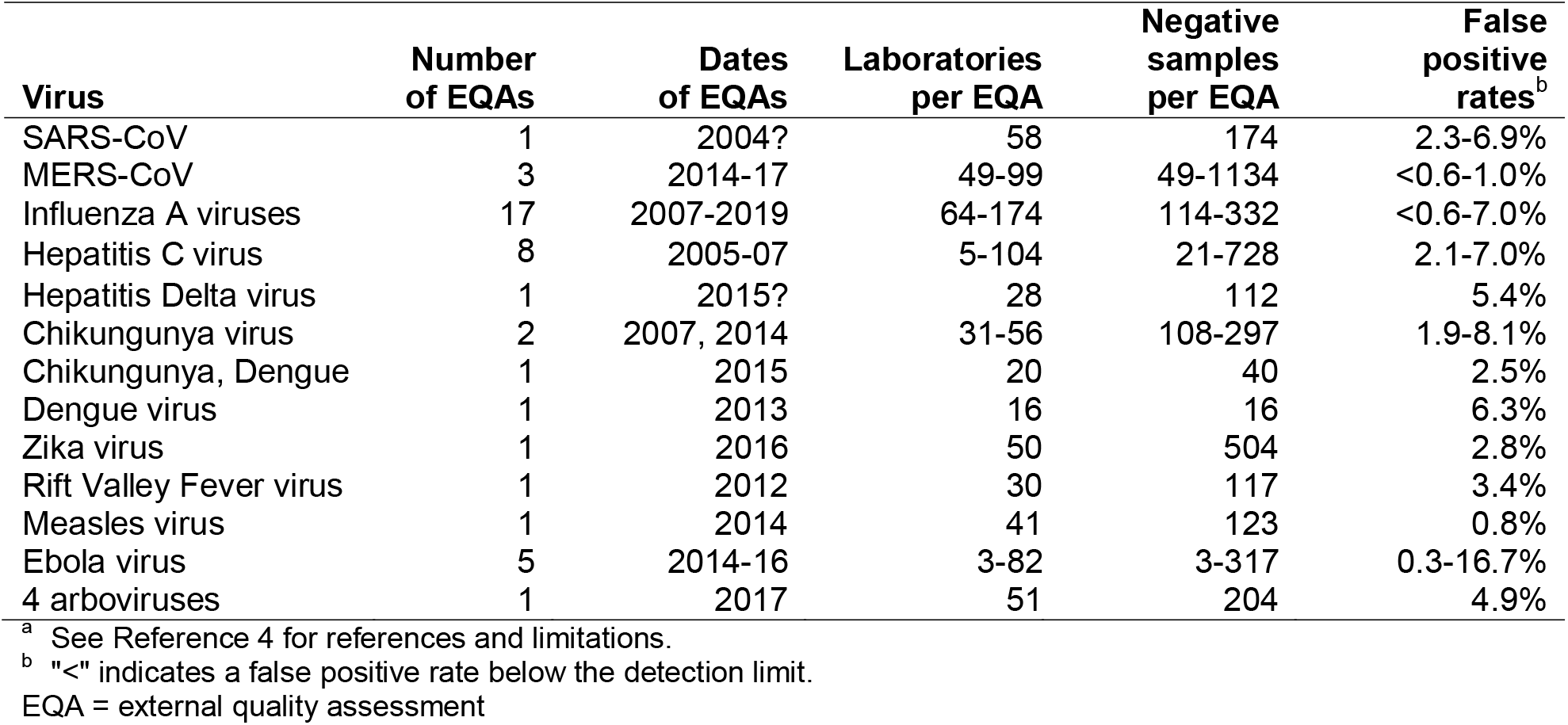
False positive rates in external quality assessments of PCR tests for RNA viruses^a^

| **Virus** | **Number of EQAs** | **Dates of EQAs** | **Laboratories per EQA** | **Negative samples per EQA** | **False positive rates<sup>b</sup>** |
| --- | --- | --- | --- | --- | --- |
| SARS-CoV | 1 | 2004? | 58 | 174 | 2.3-6.9% |
| MERS-CoV | 3 | 2014-17 | 49-99 | 49-1134 | <0.6-1.0% |
| Influenza A viruses | 17 | 2007-2019 | 64-174 | 114-332 | <0.6-7.0% |
| Hepatitis C virus | 8 | 2005-07 | 5-104 | 21-728 | 2.1-7.0% |
| Hepatitis Delta virus | 1 | 2015? | 28 | 112 | 5.4% |
| Chikungunya virus | 2 | 2007, 2014 | 31-56 | 108-297 | 1.9-8.1% |
| Chikungunya, Dengue | 1 | 2015 | 20 | 40 | 2.5% |
| Dengue virus | 1 | 2013 | 16 | 16 | 6.3% |
| Zika virus | 1 | 2016 | 50 | 504 | 2.8% |
| Rift Valley Fever virus | 1 | 2012 | 30 | 117 | 3.4% |
| Measles virus | 1 | 2014 | 41 | 123 | 0.8% |
| Ebola virus | 5 | 2014-16 | 3-82 | 3-317 | 0.3-16.7% |
| 4 arboviruses | 1 | 2017 | 51 | 204 | 4.9% |

<sup>a</sup> See Reference 4 for references and limitations.
<sup>b</sup> "<" indicates a false positive rate below the detection limit.
EQA = external quality assessment

## At low prevalence, the reliability of positive results declines

Figure 1 shows that even a false positive rate of 0.3% (the lowest rate from studies in real-world settings) can greatly reduce the reliability of test results. At that rate, in countries with a low test positivity rate, overly broad testing has produced results that are too unreliable to be useful (toward the right side of panel A, which shows measures of reliability calculated from countries’ cumulative test data). Reliability measures calculated from daily test data contrast the time course in Italy (in Panel B), which suffered a catastrophic outbreak, with that in South Korea (Panel C), which avoided one. These calculations show that in South Korea after April 20th most of the positive test results in asymptomatic individuals could have been false positives, even as the country continued to conduct over 6,000 tests a day.

**Figure 1.**
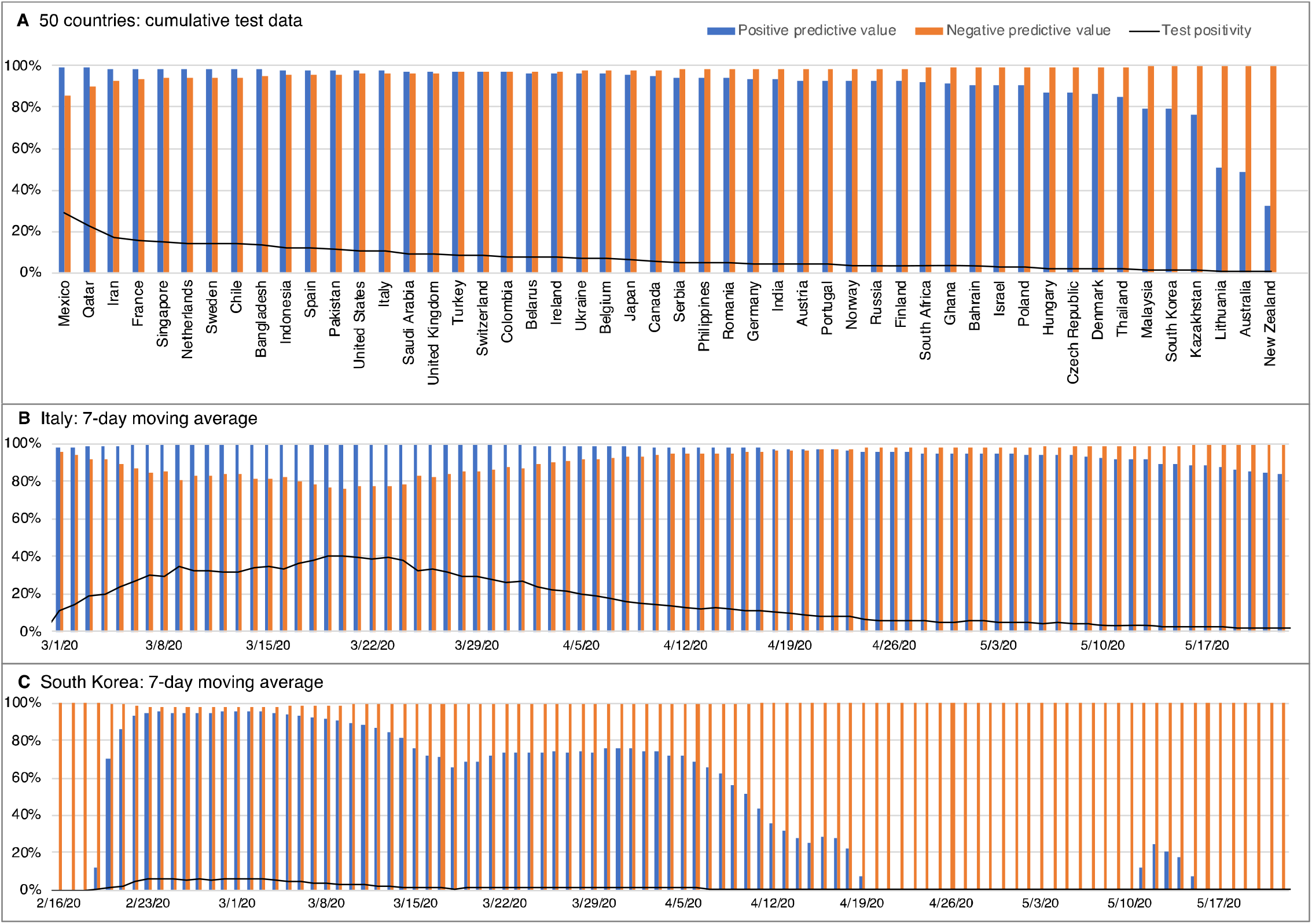
Reliability of SARS-CoV-2 test results in different countries. Positive predictive value (the probability that a positive result is true) and negative predictive value (the probability that a negative result is true) calculated with a false negative rate of 26% (midpoint of published estimates of 0-52%)^4^ and a false positive rate of 0.3%. (A) Results for the 50 countries with the greatest reported number of tests based on cumulative test data through 24 May 2020. Countries arranged left to right in order of decreasing test positivity. (B, C) Reliability trajectories based on the previous-7-day moving average, showing a country with (Italy) and without (South Korea) a major outbreak. Cumulative test data are from Our World in Data (https://github.com/owid/covid-19-data/tree/master/public/data/ accessed 24 May 2020). Daily test data are from the Italian Ministry of Health (http://www.salute.gov.it/portale/nuovocoronavirus/archivioNotizieNuovoCoronavirus.jsp?lingua=italiano&menu=notizie&p=dalministero&area=nuovocoronavirus&notizie.page=0 accessed 24 May 2020) and the South Korean Centers for Disease Control and Prevention (https://www.cdc.go.kr/board/board.es?mid=&bid=0030 accessed 24 May 2020).

The reliability of positive results falls to near zero when the test positivity rate approaches the false positive rate. However, even with positivities up to ten times the false positive rate, a significant proportion of positive results will be false. For example, with a false positive rate of 0.3% and a test positivity rate of 1% nearly 1 in 3 positive results will be false, and with a positivity rate of 3% nearly 1 in 10 will be false. Most of these false-positive individuals would likely be asymptomatic, which could at least partially explain the reports of large numbers of asymptomatic carriers of SARS-CoV-2.

Public health authorities often state that positive results from SARS-CoV-2 tests are more trustworthy than negative results.^1-4^ However, over a wide range of likely scenarios, the opposite is true: for example, in figure 1 wherever the blue columns (positive predictive values) are lower than the orange columns (negative predictive values), positive results are more likely to be wrong than are negative results. This is because the false positive rate affects samples from uninfected people, while the false negative rate affects samples from people that are infected. When prevalence is low, there are many more uninfected than infected people, so even a low false positive rate can have a larger effect than a high false negative rate.

## Sources of false positives

As with other PCR-based diagnostic tests, most false positives in SARS-CoV-2 tests are probably due to contamination, derived from such sources as positive samples, positive controls, contaminated reagents, or infected workers.^4 12^ Massive amplification of nucleic acids makes PCR-based assays highly sensitive, but also highly vulnerable to minute levels of contamination which can produce false positives that are indistinguishable from true positives. Even the most highly-regarded laboratories struggle to avoid contamination problems when using PCR, and sometimes fail.^31 32^ False positives can also be produced by sample mix-ups, software problems or data errors.^4^

## Impacts from false positives

Considerable attention has been paid to *false negative rates* in SARS-CoV-2 PCR testing^4^ and to false positive rates in *antibody* testing,^33^ but there has been little discussion in the scientific or medical literature of false positive rates in SARS-CoV-2 PCR testing.^4^ Failing to anticipate and correct for false positive results has numerous clinical and case management consequences. These include waste of personal protective equipment, waste of human resources in contact tracing,^35^ unnecessary delays in surgical procedures,^25 35^ prolongation of hospital stays,^25 35^ and potentially dangerous sequestering of uninfected individuals with infected individuals.^25 32^ A false positive test result can impede a correct diagnosis, delaying or depriving patients of appropriate treatment. False-positive patients introduce noise into clinical observations, which may hinder the development of improved medical care based on clinical experience. False-positive individuals or their close contacts could be subjected to medically inappropriate therapies,^34^ including prophylactic or antiviral medications and antibody therapy. Individuals that have falsely tested positive may be less likely to avoid future exposure to infected individuals, believing they have immunity, and for the same reason may not seek vaccination when it becomes available. Clinical trials could lose statistical power by unwittingly enrolling false-positive individuals, who would be exposed to potentially harmful side effects without any mitigating potential for benefit. False positives also distort the estimates of an array of epidemiological statistics that affect policy decisions including the asymptomatic ratio, prevalence estimates, and hospitalization and death rates, as well as many modeling studies.

## Fixing the problem

The impact of false positives in SARS-CoV-2 testing would be somewhat mitigated by merely increasing the awareness of false positives. This would introduce an appropriate note of caution into clinical and management decisions where patients might be harmed if not already infected, and would promote the inclusion of reasonable estimates of false positive rates into analyses of test data, substantially changing results in some cases. These would be helped by improved estimates of false positive rates, either from external quality assessments designed to realistically estimate false positive rates, or retrospective confirmation of PCR results with serological tests.

But more importantly, we should reduce false positive rates. Long-term, this can be done by investigating and improving laboratory and sampling practices. Shorter-term solutions, all of which involve tradeoffs between specificity and sensitivity, include raising the criteria for positive results in PCR tests by lowering the cutoff known as the maximum threshold cycle (Ct), or by selecting tests with primer-probe sets that are less sensitive, which would reduce false positives that result from a low-concentration contamination. Pooled sampling also reduces false positives. However, a simpler and immediately available approach is to check positive results with additional tests, at least when prevalence is low, such as in the mass-testing of asymptomatic individuals. In such circumstances, requiring two independent positive test results to diagnose an individual as infected greatly reduces the effective false positive rate at the cost of a minimal, often insignificant, increase in the false negative rate.

There is evidence that this works. This past spring, the provincial government of Ontario, Canada decided to test all residents and staff at long-term care homes. Medical officers overseeing three counties, anticipating the possibility of false positives, retested all positive results. Eight specimens initially tested positive, from eight asymptomatic residents and staff at eight separate homes. The individuals and their families were informed of the results but told that they were tentative pending confirmation, and the eight homes were put on lockdown in the interim. Second aliquots taken from the eight original samples all tested negative, as did second and third swabs from the eight individuals. This additional testing increased the total number of tests used by 0.5%. The medical officers concluded that the initial positive results were false, informed the eight individuals, and ended the lockdowns (I. Arra, personal communication; D. Colby, personal communication).

Now imagine what would have happened without retesting. The initial results would be accepted as proof of infection, the individuals would be told they have a disease that stands a good chance of killing them in short order, and the 558 residents of the eight homes would be put in lockdown, restricted to their rooms without visitors or activities, for 14 days. Residents, staff and their families would be subject to greater and longer-endured levels of anxiety, with potentially greater physical and mental health impacts on the isolated residents. During that time, the residents with false positive results would be attended only by staff in full PPE, causing unnecessary consumption of these supplies and further isolation of the residents. Unnecessary contact tracing would be conducted for any of the eight that had outside contacts, potentially resulting in additional, unnecessary tests. In some localities the eight false positives would require two additional, negative tests in order to leave isolation. In this case there were no infected individuals in the homes for the false-positive residents to be sequestered with; however, in some localities they would be transferred to a common facility with infected individuals, significantly elevating the risk of infection and, for the elderly or vulnerable, the risk of death.

Like all tests, PCR-based assays are subject to error that includes both false negative *and* false positive results. A successful testing program must understand the error rates of both and use tests appropriately. While SARS-CoV-2 testing to date has clearly missed the mark, we can course-correct: we can reassess plans for mass-testing using realistic estimates of false-positive rates, reconsider the conclusions of studies that implicitly assumed a zero false positive rate, and reduce misdiagnoses and statistical miscounts by checking positive results with follow-up tests, especially in asymptomatic persons and in areas where test positivity is low. In the interim, where testing has been conducted without regard to symptoms or exposure—notably in certain localities, congregate-living facilities, workplaces and sports leagues—positive results in healthy individuals should be considered doubtful unless confirmed by a second test.

## Data Availability

All relevant data are included in the text or are available from the corresponding author on request.

## Contributors

ANC is a marine biologist with expertise in biological invasions; this analysis grew out of previous work on false positives in PCR-based assays used in environmental monitoring. BK is a clinical Ob/Gyn academic whose research has ranged from menopause to ovarian cancer. MGM has used PCR-based analyses to investigate plant diseases. ANC designed and conducted the analyses with input from MGM; ANC and BK collaborated on the review of clinical implications; and all authors participated in writing and editing the report. ANC is the guarantor. We thank Elisa Liberati, Dominic Chow, Thomas Taylor, James Carlton, Richard Norgaard, Bruno Pernet, Ian Arra and David Colby for helpful comments and other assistance. This paper is dedicated to the memory of Kirk Smith, who kindly reviewed the manuscript and offered insightful advice a few weeks before his untimely death. All data is available in the manuscript or from the authors on request. No funding was received for this work. The authors declare no conflicts of interest.

## Notes

### Competing Interest Statement

The authors have declared no competing interest.

### Funding Statement

The was no funding for this research.

### Author Declarations

All necessary approvals have been obtained.

### Summary of Updates

The material in the text and the figure have been updated; the figure now includes data for fewer countries, to improve readability. The material previously included in the Supplemental Material file has been updated and partially rewritten; that file has been deposited in an online respository referenced with its URL as a citation in the text.

